# Neurocognitive Dysfunction in Children and Adolescents with Mental Illness

**DOI:** 10.1101/2023.02.23.23286376

**Authors:** Sean Nesamoney, Rachel A. Hilton, Leonardo Tozzi, Leanne M. Williams

## Abstract

**Introduction:** Robust evidence from adult samples indicates that neurocognitive dysfunction is a hallmark of many mental illnesses, contributing to the loss of daily function and quality of life that these illnesses cause. However, it is still unclear whether neurocognitive deficits associated with mental illnesses begin to manifest well before adulthood. The current study addresses this gap by evaluating neurocognitive function in four groups of children and adolescents with different mental illnesses compared to their matched healthy peers.

**Methods:** We evaluated the neurocognitive performance of four samples of youth diagnosed with ADHD (N=343), Anorexia (N=40), First-onset psychosis (N=25), and Conversion Disorder (N=56) with age-matched healthy controls. Performance was assessed using an objective assessment battery designed for use across diagnoses and settings and validated for its correlations with underlying brain structure and function. The resulting analyses assessed accuracy and reaction time performance for neurocognitive domains well established in the adult literature, such as cognitive flexibility, executive function, response inhibition, verbal fluency, verbal memory, visual memory, sustained attention, and working memory. Clinical and healthy group performance was compared using non-parametric testing.

**Results:** Distinct profiles of neurocognitive dysfunction were detected for each diagnosis. Particularly, children and adolescents with ADHD diffusely performed worse than their healthy counterparts, with exceptional impairment in working memory. Children and adolescents with anorexia displayed more specific impairments limited to response inhibition and verbal memory. While youth with ADHD had the most cognitive domains affected, youth with first-onset psychosis displayed the most severe impairments compared to healthy controls. Finally, deficits in conversion disorder were limited to cognitive flexibility, executive function, decision making, response inhibition, and working memory. These findings suggest that neurocognitive impairment in mental illness is transdiagnostic and can be detected as early as childhood or adolescence with standardized computerized testing.

## Introduction

Cognitive dysfunction is a diagnostic feature of multiple mental illnesses among adults and youth, most notably attention-deficit hyperactivity disorder (ADHD). Patients’ performance on neuropsychological tests can be used to quantify cognitive dysfunction. These tests involve performing tasks and can provide objective measurements of cognitive domains such as set-shifting, response inhibition, working memory, fluency, planning, verbal memory, non-verbal memory, processing speed, attention, and visuospatial function ^1^.

However, there is also strong evidence of cognitive dysfunction in disorders not traditionally defined by changes in cognition. A 2021 systematic review of meta-analyses on the topic ^2^ compared cognitive performance in 97 samples of patients suffering from various psychiatric disorders to cognitive performance in healthy controls. Results indicated a significant impairment in at least one cognitive domain in patients with autism spectrum disorder (ASD), bipolar disorder (BD), eating disorders, depression, schizoaffective disorder, obsessive-compulsive disorder (OCD), personality disorders, post-traumatic stress disorder (PTSD), schizophrenia, substance use disorders (SUD), and Tourette’s syndrome, with effect sizes ranging from 0.4 to 0.6. Disorders characterized by psychotic symptoms, such as schizophrenia and BD, showed the largest deficits in cognitive performance. Conversely, eating disorders and SUD showed the smallest cognitive deficits. Other recent investigations have produced similar results ^3–8^.

Overall, these results suggest that cognitive deficits are a transdiagnostic feature of mental illness, i.e., a feature that cuts across traditional diagnostic boundaries. Therefore, they are not limited to disorders using cognitive dysfunction as part of their diagnostic criteria. To date, studies have largely investigated cognitive impairments in adults with mental illness, despite the majority of mental illnesses emerging in childhood and adolescence ^9–12^. Mental illnesses are the leading cause of disability in children and adolescents, affecting an estimated 13.4% of youth worldwide ^13,14^. It is unclear if cognitive deficits associated with mental illness begin to manifest before adulthood, although preliminary evidence suggests they might ^15–19^.

The current study addressed this gap by comparing cognitive performance in patients aged six to eighteen diagnosed with ADHD, anorexia, first-onset psychosis, and conversion disorder with the cognitive performance of age-matched healthy controls. Cognitive performance was assessed using a standardized web-based cognitive battery ^1^, thus enabling us to quantify cognitive deficits in each clinical diagnosis consistently. We focused on the cognitive domains of cognitive flexibility, executive function, decision making, response inhibition, verbal fluency, verbal memory, visual memory, sustained attention, and working memory to align with previous adult literature.

We hypothesized that youth patients suffering from the included diagnoses would display cognitive deficits compared to age-matched controls and that findings would mirror previous findings in adult clinical populations.

## Methods

### Participants

The data used in the present work was downloaded from Stanford BRAINnet, a large database for mental health research (www.stanfordbrainnet.com). In particular, the current sample was recruited at the University of Sydney through advertising and self-referral. Inclusion criteria were age between 6 and 18 years old, diagnosis of ADHD, anorexia, first-onset psychosis, or conversion disorder (clinical groups) or no psychiatric diagnosis (controls), capacity to undergo a computerized test, reading at Year 5 level (equivalent to fifth grade in the United States), normal (or corrected to normal) vision, and ability to use a keyboard. The protocol received an independent ethics committee or institutional review board approval before recruitment of participants. All participants signed and dated an approved informed consent form. Where participants consented, their data were made available for open sharing in Stanford BRAINnet. All research complies with the Code of Ethics of the World Medical Association (Declaration of Helsinki).

Diagnosis of clinical groups was confirmed by consensus from clinicians using the following criteria and scales. For ADHD: Structured Clinical Interview (DSM-IV) and Conners’ Rating Scale ^20^, for details, see ^21^. For anorexia: Structured Clinical Interview (DSM-IV) and physical conditions, for details see ^22^. For first-onset psychosis: Structured Clinical Interview (DSM-IV) and Positive and Negative Symptoms Scale ^23^, for details see ^24^. For conversion disorder: Structured Clinical Interview (DSM-IV), for details see ^25^.

For each clinical group, an age and sex-matched sample of healthy individuals was recruited as a control group by the same site.

A summary of the demographic characteristics of the participants is given in Table 1.

**Table 1:** Study demographics. Age and sex compositions are shown for each clinical group and their matched health controls. F=female, M=male, SD= standard deviation

|  | **ADHD** |  | **Anorexia** |  | **First-onset**<br>**Psychosis** |  | **Conversion**<br>**disorder** |  |
| --- | --- | --- | --- | --- | --- | --- | --- | --- |
|  | Clinical | Control | Clinical | Control | Clinical | Control | Clinical | Control |
| **N** | 343 | 343 | 40 | 41 | 25 | 42 | 56 | 57 |
| **Age**<br>**range**<br>**(years)** | 6-18 | 6-18 | 12-18 | 12-18 | 11-18 | 11-18 | 8-18 | 8-18 |
| **Age**<br>**mean and**<br>**SD**<br>**(years)** | 11.78±<br>3.20 | 11.87±<br>3.08 | 15.23±<br>1.61 | 15.25±<br>1.58 | 16.23±<br>1.63 | 16.19±<br>1.37 | 13.49±<br>2.12 | 13.60±<br>2.31 |
| **Sex** | F = 76<br>(22.2%);<br>M = 267<br>(77.8%) | F = 76<br>(22.2%);<br>M = 267<br>(77.8%) | F = 40<br>(100%);<br>M=0<br>(0%) | F = 41<br>(100%);<br>M=0<br>(0%) | F = 10<br>(40%);<br>M = 15<br>(60%) | F = 12<br>(28.6%);<br>M = 30<br>(71.4%) | F = 40<br>(71.4%);<br>M = 16<br>(28.6%) | F = 41<br>(71.9%);<br>M = 16<br>(28.1%) |

### Neurocognitive assessments

Neurocognitive data was collected using IntegNeuro, a standardized battery validated in healthy and clinical populations^1^. IntegNeuro has established test-retest reliability and has been validated for its correlation with brain structure and function. To compare results to findings from a recent systematic review of meta-analyses on cognitive dysfunction in adult psychiatric patients, we focused our analyses on the following WebNeuro domains, cognitive flexibility, executive function, decision making, response inhibition, verbal fluency, verbal memory, visual memory, sustained attention, and working memory ^2^. For each of these domains, the cognitive tasks used are outlined below and displayed in Table 2.

**Table 2:** Cognitive domains assessed with corresponding tasks and measures.

| Domain | Test | Measure | Equivalent Neuropsychological test |
| --- | --- | --- | --- |
| Cognitive Flexibility | Switching of Attention | Errors (Part 1, 2) | Trails A, B |
|  |  | Completion Time (Part 1, 2) |  |
| Executive Function | Maze | Completion time | Austin Maze |
|  |  | Errors |  |
| Decision Making | Choice Reaction | Reaction time |  |
| Response Inhibition | GoNoGo | Reaction time |  |
|  |  | False Alarms |  |
|  | Verbal Interference | False Misses |  |
|  |  | Accuracy (Ignore Color, Ignore Word) | Stroop |
| Verbal Fluency | Verbal Fluency | Number of Words Generated | F,A,S |
|  | Semantic Fluency | Number of Animals Named | Semantic Fluency |
| Verbal Memory | Immediate | Accuracy | California Verbal Learning Test |
|  | Short Delay | Accuracy |  |
|  | Long Delay | Accuracy |  |
| Visual Memory | Span of Visual Memory | Accuracy | Corsi Blocks |
| Working Memory | Digit Span | Accuracy (Forward, Reverse) |  |
| Sustained Attention | Continuous Performance | Reaction Time | Connors CPT, TOVA |
|  |  | False Positive Errors |  |
|  |  | False Negative Errors |  |

#### Cognitive Flexibility

A switching of attention task was used to assess this domain. This task consists of two parts. In the first part, the participant is presented with a pattern of 25 numbers in circles and asked to touch them in ascending numerical sequence (i.e., 1 2 3 …). In the second part, the participant is presented with a pattern of 13 numbers (1–13) and 12 letters (A–L) on the screen and is required to touch numbers and letters alternatively in ascending sequence (i.e., 1 A 2 B 3 C …). This part requires the participant to switch attention between mental tasks, in this case, number and letter sequence checking. Task outputs for analysis included the time to complete each of the two parts and the number of errors in each of the two parts.

#### Executive Function

A maze task was used to assess this domain. The participant is presented with a grid of circles on the computer screen. The object of the task is to identify the hidden path through the grid, from the beginning point at the bottom of the grid to the end point at the top. The participant can navigate around the grid by pressing arrow keys (up, down, left, right). A total of 24 consecutive correct moves are required to complete the maze. After each move, the participant receives feedback on whether it was incorrect or correct. The purpose of the task is, therefore, to assess how quickly the participant learns the route through the maze and their ability to remember that route. The outputs of this task used for analysis included the time to complete the maze and the number of errors.

#### Decision Making

A choice reaction time task was used to assess this domain. Participants are required to attend to the computer screen as one of four target circles is illuminated in pseudorandom sequence over a series of trials. For each trial, the participant is required to click on the illuminated circle as quickly as possible. Twenty trials are administered with a random delay between trials of 2–4 seconds. Mean reaction time across trials was used for analysis.

#### Response Inhibition

Verbal interference and go-no-go tasks were used to assess this domain. In the verbal interference task, the participant is presented with colored words, one at a time. Each word is drawn from the following lowercase words: red, yellow, green, and blue. The color of each word is drawn from the following set of colors: red, yellow, green, and blue. Below each colored word is a response pad with the four possible words displayed in black and in a fixed format. The test has two parts. In the first part, the participant is required to identify the name of each word as quickly as possible after it is presented on the screen. In the second part, the participant is required to name the color of each word as quickly as possible. Outputs for the resulting analysis included the number of correct responses in each of the two parts. In the go-no-go task, the word PRESS is frequently presented in green (go) and infrequently in red (no-go). The participant is required to inhibit keypress responses on red. This task measures target detection rate, response time, errors of commission, and omission. The outputs of this task used for analysis included average response time, number of responses in no-go trials, and number of non-responses in go trials.

#### Verbal Fluency

Letter and animal fluency tasks were used to assess this domain. In the letter fluency task, participants were required to say words that began with the letters F, A, and S. Sixty seconds were allowed for each letter, and proper nouns were not allowed. Responses were recorded via a microphone and hand-scored. Outputs included the average number of words generated across letters. In the animal fluency task, participants were required to name animals as quickly as possible for 60 seconds, and outputs included the number of animals named.

#### Verbal Memory

A verbal list-learning task was used to assess verbal memory. The participants were read four times a list of 12 concrete English words, which they were asked to memorize. On each of the four immediate recall trials, the participant was required to recall as many words as possible. The participant was then presented with a list of distractor words and asked to remember them. Immediately following this, the participant was asked to recall the 12 words from the original list (short delay recall trial). A long delay recall trial was completed approximately 20 min later after a few intervening tasks. A recognition trial was then completed after the long delay recall trial. The outputs of this task used for analysis were the number of correct words recalled in the four immediate recall trials, the number of correct words recalled in the long delay recall trial, and the number of correct words recalled in the short delay recall trial.

#### Visual Memory

A span of visual memory task was used to assess this domain. Participants were presented with squares arranged in a random pattern on the computer screen. The squares are highlighted in sequential order on each trial. Participants were required to repeat the order in which the squares were highlighted in both forward and reverse order. Outputs included the number of correct responses in forward and reverse trials.

#### Working Memory

A digit span task was used to assess this domain. Participants repeat a span of 2 to 9 numbers, and behavior is quantified as maximum span.

#### Sustained Attention

A continuous performance task was used to assess this domain. A series of letters (B, C, D, or G) were presented to the participant on the computer screen (for 200 msec), separated by an interval of 2.5 sec. The participant had to respond if the same letter appears twice in a row. Outputs included the average response time, the standard deviation of the response time, the number of incorrect responses, and the number of missed responses.

### Reference of cognitive performance to a healthy norm

Statistical analyses used the corrected version of the 24 task outputs described above, which is an output provided by WebNeuro. These values are obtained by expressing the task outputs of each participant as scores based on their deviation from a reference healthy population, matched by sex and age ^26^.

### Statistical analysis

All analyses were conducted utilizing the Statistical Product and Service Solutions (SPSS) Statistics version 28.0.1.1 (14) for Mac. We compared the cognitive variables of each clinical group (ADHD, anorexia, first-onset psychosis, and conversion disorder) with those of their matched healthy control group. Mann-Whitney U tests were used to evaluate for any differences in cognition between clinical patients and controls. Mann-Whitney U tests were employed due to variables following a non-normal distribution (e.g. count distribution for correct responses). False-discovery rate (FDR) corrections accounted for the testing of multiple cognitive variables for each clinical group and minimized false positive results ^27^. Values were considered statistically significant when the FDR corrected p-value (pFDR) was less than 0.05. To determine the effect size of these differences, Cohen’s d values were calculated to determine effect sizes of found differences.

## Results

### Cognitive assessments

Table 3 summarizes the performance of each group of participants along with the results of statistical tests comparing each group of clinical participants with controls. Figure 1 presents a summary of cognitive impairments across the included diagnoses.

**Table 3:** Cognitive deficits in patients with attention-deficit hyperactivity disorder (ADHD), Anorexia, First-Onset Psychosis, and Conversion Disorder. Clinical groups were compared to matched controls using Mann-Whitney U tests. To account for multiple comparisons, the p-values obtained from the test were corrected using false-discovery rate (PFDR). Bolded values met statistical significance (PFDR ≤ 0.05).

|  |  |  | ADHD > Controls |  | Anorexia > Controls |  | FEP > Controls |  | Conversion > Controls |  |
| --- | --- | --- | --- | --- | --- | --- | --- | --- | --- | --- |
| Domain | Test | Measure | d | $p_{FDR}$ | d | $p_{FDR}$ | d | $p_{FDR}$ | d | $p_{FDR}$ |
| Cognitive Flexibility | Switching of Attention | Part 1 Errors | -0.287 | **<0.001** | 0.011 | 0.998 | 0.115 | 0.712 | 0.341 | 0.168 |
|  |  | Part 1 Completion Time | 0.196 | **0.016** | -0.305 | 0.367 | 0.474 | 0.092 | 0.540 | **0.034** |
|  |  | Part 2 Errors | 0.517 | **<0.001** | -0.390 | 0.250 | 0.094 | 0.754 | 0.098 | 0.690 |
|  |  | Part 2 Completion Time | 0.234 | **0.004** | -0.431 | 0.250 | 0.550 | 0.053 | 0.359 | 0.168 |
| Executive Function | Maze | Completion time | 0.447 | **<0.001** | 0.327 | 0.367 | -0.013 | 0.960 | 0.549 | 0.144 |
|  |  | Errors | 0.510 | **<0.001** | 0.078 | 0.842 | -0.202 | 0.532 | 1.006 | **<0.001** |
| Decision Making | Choice Reaction | Reaction Time | 0.242 | **0.004** | 0.130 | 0.725 | 0.626 | **0.034** | 0.495 | **0.048** |
| Response Inhibition | GoNoGo | Reaction Time | 0.360 | **<0.001** | 0.581 | 0.125 | 0.540 | 0.061 | -0.562 | **0.034** |
|  |  | False Alarms | 0.368 | **<0.001** | -0.156 | 0.725 | 0.772 | **0.012** | 0.181 | 0.490 |
|  |  | False Misses | 0.615 | **<0.001** | 1.235 | **<0.001** | 1.229 | **<0.001** | -0.534 | **0.034** |
|  | Verbal Interference | Accuracy (Ignore Color) | -0.301 | **<0.001** | 0.017 | 0.998 | -0.730 | **0.012** | 0.202 | 0.490 |
|  |  | Accuracy (Ignore Words) | -0.141 | 0.078 | 0.127 | 0.725 | -0.705 | **0.015** | 0.072 | 0.773 |
| Verbal Fluency | Verbal Fluency | Number of Words Generated | -0.372 | **<0.001** | 0.526 | 0.125 | -0.783 | **0.012** | -0.191 | 0.490 |
|  | Semantic Fluency | Number of Animals Named | -0.479 | **<0.001** | 0.086 | 0.842 | -1.101 | **<0.001** | -0.034 | 0.906 |
| Verbal Memory | Immediate | Accuracy | -0.583 | **<0.001** | 0.146 | 0.725 | -1.419 | **<0.001** | -0.261 | 0.397 |
|  | Short Delay | Accuracy | -0.474 | **<0.001** | -0.827 | **<0.001** | -1.382 | **<0.001** | 0.136 | 0.576 |
|  | Long Delay | Accuracy | -0.386 | **<0.001** | -0.351 | 0.360 | -1.500 | **<0.001** | 0.173 | 0.507 |
| Visual Memory | Span of Visual Memory | Accuracy | -0.470 | **<0.001** | 0.142 | 1.000 | 0.060 | 0.827 | 0.156 | 0.515 |
| Working Memory | Digit Span | Accuracy (forward) | -0.469 | **<0.001** | 0.143 | 0.725 | -0.731 | **0.012** | -0.735 | **<0.001** |
|  |  | Accuracy (reverse). | -0.346 | **<0.001** | 0.295 | 0.367 | -0.294 | 0.315 | -0.493 | **0.048** |
| Sustained Attention | Continuous Performance | Reaction Time | 0.364 | **<0.001** | 0.546 | 0.125 | 0.606 | **0.037** | -0.187 | 0.490 |
|  |  | False Positive Errors | 0.671 | **<0.001** | 0.381 | 0.300 | 0.247 | 0.367 | 0.186 | 0.490 |
|  |  | False Negative Errors | 0.555 | **<0.001** | 0.306 | 0.367 | 1.166 | **<0.001** | 0.184 | 0.490 |

**Figure 1:**
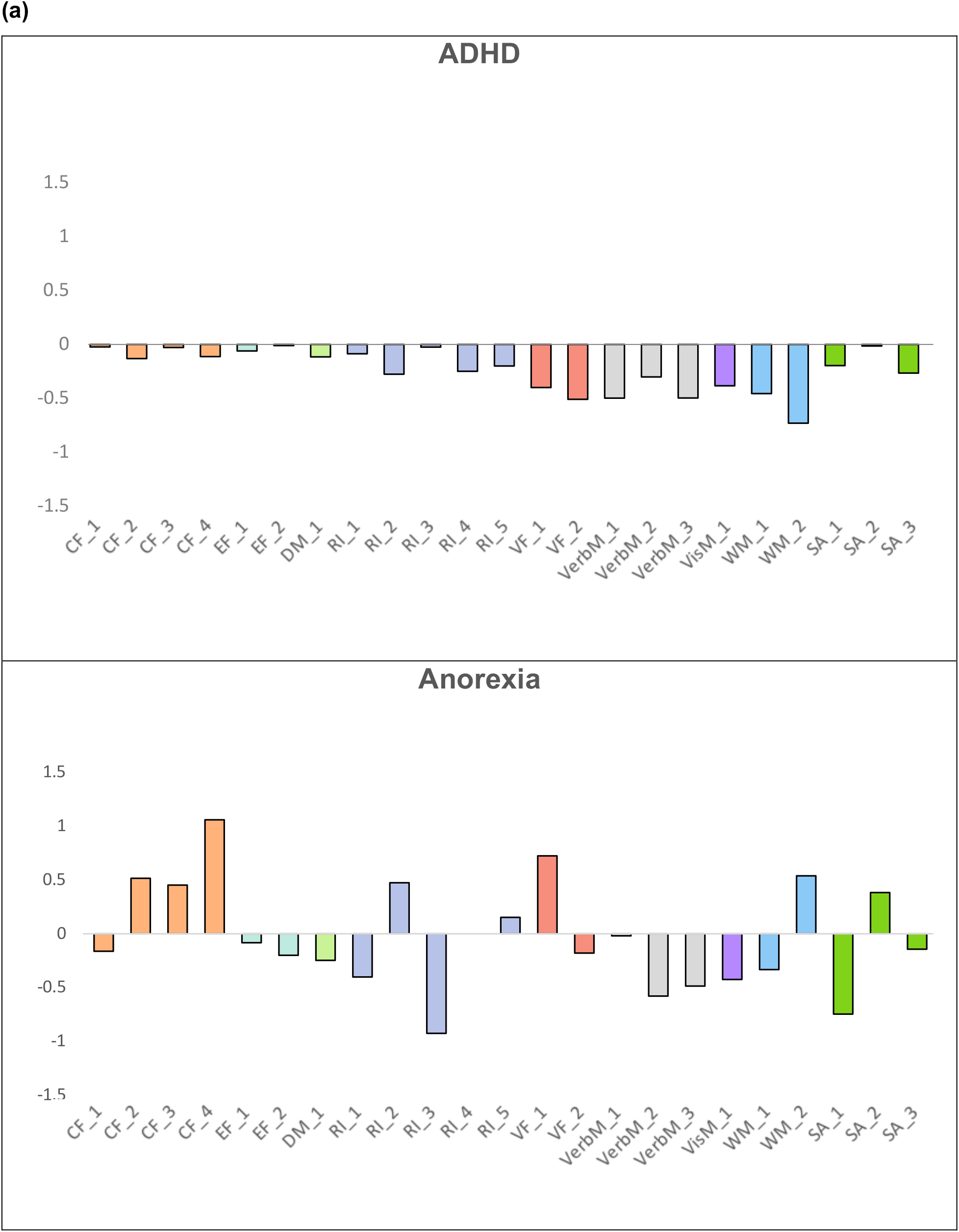
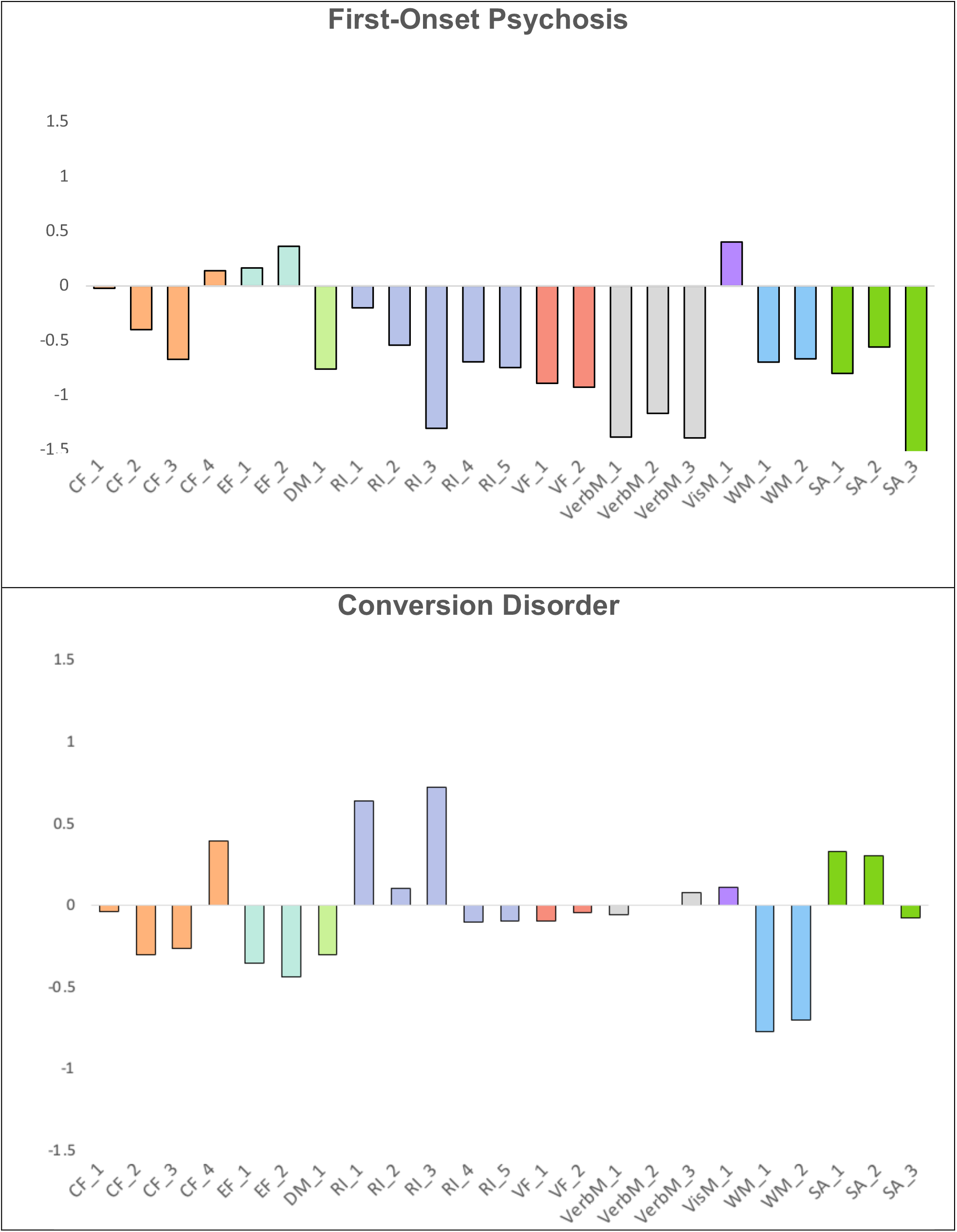
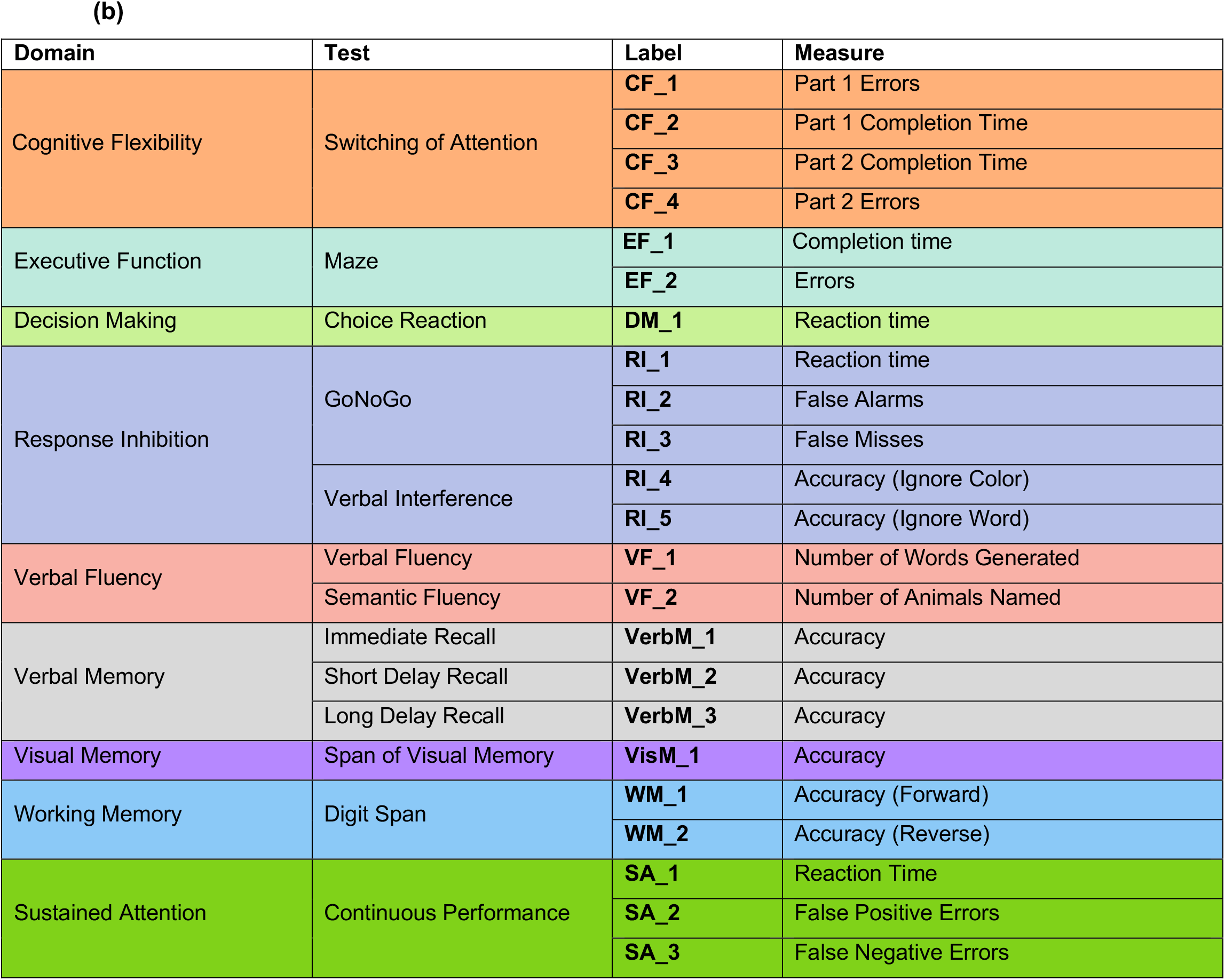
Summary of cognitive deficits across diagnostic groups. (a) Values represent median scores for each task. To assist interpretability, some raw scores have been inverted so that positive values indicate improved performance on tests compared to healthy controls and negative values represent worse performance on tests compared to healthy controls. (b) Legend explaining task labeling.

Consistent with our hypotheses, all clinical groups showed a significant impairment of at least one cognitive function measure compared to healthy matched controls. The following sections outline these deficits in detail.

#### ADHD

Patients with ADHD performed worse than controls across all measures (Table 3). Deficits within working memory were particularly strong, as shown by ADHD patients taking much longer on average to complete a continuous performance task (U=35355, d=0.364, pFDR<0.001) with a higher error (U=28289, d=0.671, pFDR<0.001) and missed response rate (U=30923, d=0.555, pFDR<0.001).

#### Anorexia

Performance differences between patients with anorexia and their healthy counterparts were largely statistically insignificant with the exception of performance in selective tasks within the response inhibition and verbal memory domains (Table 3). In the go-no-go task, patients had a higher number of non-responses in go trials (U=249, d=1.24, pFDR<0.001). In the verbal memory task, they were able to recall fewer words in the short delay recall condition (U=389, d=-0.827, p<0.001).

#### First-onset psychosis

Patients with first-onset psychosis performed worse than controls across all cognitive domains except cognitive flexibility, executive functioning, and visual memory (all other pFDR<0.05, Table 3). Impairments in verbal memory were most pronounced compared to their healthy counterparts, with effect sizes ranging from -1.38 to -1.50 (all pFDR <0.001). Patients also displayed striking deficits in response inhibition and working memory (Table 3). Regarding response inhibition, patients with psychosis specifically performed worse responding inappropriately in “no-go” trials (U=245, d=0.77, pFDR=0.012) and missing responses in “go” trials (U=156, d=1.23, pFDR<0.001). During tasks challenging working memory, patients had significantly fewer correct responses during a forward digit span task (U=289, d=-0.73, pFDR=0.012). Within the sustained attention domain, clinical patients had longer average reaction time (U=298, d=0.61, pFDR=0.037), and higher missed response rate (U=177, d=1.17, pFDR<0.001) during a continuous performance task. Interestingly, differences in switching of attention and maze task performances did not meet statistical significance (all pFDR >0.05).

#### Conversion disorder

Patients with conversion disorder showed selective deficits in cognitive flexibility, decision making, executive function, response inhibition, and working memory (Table 3). Patients with conversion disorder had longer completion times than healthy controls in multiple tasks. Longer completion/reaction times were seen in the attention-switching task (U=973 d=0.54, pFDR=0.03), choice reaction task (U=989, d=0.48, pFDR=0.05), and go-no-go task (U=979 d=-0.56, pFDR=0.03). In the go-no-go task, patients missed more responses in “go” trials (U=1001, d=-0.53, pFDR=0.03). Finally, during the digit span task, patients provided fewer correct responses during both the forward (U=801 d=-0.74, pFDR<.001) and reverse task (U=970 d=-0.49, pFDR=0.05) versions.

## Discussion

Previous investigations of adults have shown that cognitive dysfunction is a key feature of mental illness regardless of specific diagnosis ^2^. We aimed to extend these findings to children and adolescents suffering from ADHD, anorexia, first-onset psychosis, and conversion disorder. Consistent with our hypotheses, all clinical groups displayed impairments within at least one cognitive domain compared to age-matched healthy controls. Our findings suggest that cognitive dysfunction exists as a transdiagnostic feature of mental illness that can be identified with standardized cognitive testing as early as childhood and adolescence.

Interestingly, each clinical group sampled displayed a distinct pattern of cognitive dysfunction. For example, children and adolescents with ADHD showed a diffuse pattern of small to moderate differences in performance with healthy controls. However, children and adolescents with ADHD displayed exceptional impairment in working memory, for which the difference in performance was large. Broad impairment found in our sample aligns with results in adult literature which indicated impairments in verbal fluency, response inhibition, working memory, reaction time, visual memory, and executive function ^28–31^.

Compared to ADHD, cognitive deficits in youth anorexia were more specific and limited to response inhibition and verbal memory. These findings slightly differ from prior adult literature. While verbal memory deficits were previously found in adults with anorexia ^32^, patterns of response inhibition were more conflicting ^33^.

Although youth with ADHD displayed impairments in the greatest number of domains, youth patients with first-onset psychosis displayed the most severe impairment of any clinical group, with effect sizes ranging from d=0.626 to d=1.50. This youth sample’s pattern and intensity of dysfunction appear consistently within adult samples ^2^.

Finally, deficits in conversion disorder were limited to cognitive flexibility, executive function, response inhibition, decision making, and working memory. Although sparse, adult literature also suggests impairments in working memory and response inhibition in these patients ^34^. Limited evidence also shows that adults with conversion disorder also suffer from executive function deficits ^34^.

The current study is subject to limitations. First, the sample analyzed features only four clinical diagnoses: ADHD, anorexia, first-onset psychosis, and conversion disorder. Future studies should extend investigations of cognitive impairments to other mental illnesses common in childhood and adolescence, such as mood and anxiety disorders. Since every diagnostic group in our sample featured some degree of cognitive impairment, we anticipate that future studies investigating other diagnoses will produce similar results. Another limitation was that we could not consider the potential confounding effects of treatment in analyses. Future studies should account for the potential impact of treatments on cognitive performance by clinical diagnosis and assess if these treatments improve cognitive deficits.

Our findings support the assertion that cognitive impairment is a pervasive feature of mental illness, even in diagnoses that do not include cognitive impairment as a core diagnostic criterion (such as anorexia). Importantly, our findings also demonstrate that cognitive impairment in mental illness can be detected as early as childhood or adolescence with standardized computerized testing. Future studies can build on this work by investigating how cognitive impairments relate to and potentially predict clinically important outcomes, such as lower educational achievement, worse quality of life, treatment response, or suicidality.

## Data Availability

The data used in the present work was downloaded from and is available within Stanford BRAINnet, a large database
for mental health research (www.stanfordbrainnet.com).

## Acknowledgements

The authors would like to extend their appreciation to the participants in the studies from which source data are drawn. The data are drawn from Stanford BRAINnet, which includes data acquired with support to LMW from the National Health and Medical Research Council (Project Grants 1004822, 457424), Australian Research Council (Discovery Projects DP120104496, DP077394, DP0345481), and a Pfizer Foundation Senior Research Fellowship.

## Ethics Approval and Consent to Participate

The Institutional Review Board approved the protocols of the studies that contributed source data. A study coordinator thoroughly explained the protocol to participants and answered any questions before they provided written informed consent to begin the study. The study was conducted according to the principles of the Declaration of Helsinki.

## Competing Interests

The authors have declared no competing interest.

## Authors’ Contribution

SN, LT, RH, and LMW developed the study; SN, LT, and RH performed the data analysis. LMH, SN, LT, and RH wrote the manuscript, and all authors reviewed and approved the manuscript.

## Notes

### Author Declarations

The data used in the present work was downloaded from Stanford BRAINnet, a large database for mental health research (www.stanfordbrainnet.com).

